# Characterization of the SARS-CoV-2 outbreak in the State of Qatar, February 28-April 18, 2020

**DOI:** 10.1101/2020.07.15.20154211

**Authors:** Hanan M. Al Kuwari, Hanan F. Abdul Rahim, Laith J. Abu Raddad, Abdul-Badi Abou-Samra, Zaina Al Kanaani, Abdullatif Al Khal, Einas Al Kuwari, Salih Al Marri, Muna Al Masalmani, Hamad Eid Al Romaihi, Sheikh Mohammed Al Thani, Peter Coyle, Ali Nizar Latif, Robert Owen, Roberto Bertollini, Adeel A. Butt

## Abstract

**Objective:** To define the epidemiologic curve of COVID-19 in Qatar, determine factors associated with severe or critical illness, and study the temporal relation between public health measures and case finding

**Design:** Epidemiologic investigation

**Setting and Participants:** All confirmed COVID-19 cases in the State of Qatar between February 28 and April 18, 2020

**Main Outcome Measures:** Number of total and daily new COVID-19 infections; demographic characteristics and comorbidity burden and severity of infection; factors associated with severe or critical illness

**Results:** Between February 28 and April 18, 2020 (11:00AM local time), 5,685 cases of COVID-19 were identified. Mean age (SD) was 35.8(12.0) years, 88.9% were male and 8.7% were Qatari nationals. Overall, 83.6% had no concomitant comorbidity, and 3.0% had 3 or more comorbidities. The overwhelming majority (90.9%) were asymptomatic or with minimal symptoms, with 2.0% having severe or critical illness. Presence of hypertension or diabetes were associated with a higher risk of severe or critical illness. Seven deaths were observed during the time interval studied. The epidemiologic curve indicated two distinct patterns of infection, a larger cluster among expatriate craft and manual workers, and a smaller one among Qatari nationals returning from abroad during the epidemic.

**Conclusion:** COVID-19 infections in Qatar started in two distinct clusters, but then became more widespread in the population through community transmission. Infections were mostly asymptomatic or with minimal symptoms and associated with very low mortality. Severe/critical illness was associated with presence of hypertension or diabetes.

Article Summary
Strengths and limitations of this study:

- National study with unified contact tracing and testing
- All testing done at a single lab, and all tests performed in the State of Qatar during the study period were included, providing a robust national estimate of the number of infected persons among those tested
- Comorbidities were retrieved from the electronic medical records using ICD-10 AM codes
- Exact geographic location and contact tracing data were not included in the current report
- It is possible that some persons still under care on the study end date may have progressed to more severe disease after that date

What is already known on this topic
As of May 3, 2020, over 3.4 million persons have been infected with SARS-CoV-2 and over 244,000 deaths have been reported in persons with COVID-19 infection. Those at higher risk of complications include persons over age 60 years and those with chronic comorbid conditions. Mortality rate varies widely among different countries, and this can be associated both to the capacity of the health system to provide effective intensive care including ventilators as well as other factors, including demographic differences. Public health measures seem effective, but there is debate on the extent to which the measures need to be aggressive or the duration for which they should be implemented.

What this study adds
This study reports on the epidemic curve in a population with a unique demographic structure, comprising an overwhelming majority of expatriates and young male craft and manual workers. This is also the first study that reports on the epidemic curve of an Arab country in the Eastern Mediterranean Region (EMR). The study also overlays major public health measures on the epidemic curve, to provide an understanding of the context in which the epidemic is progressing. Patients with confirmed COVID-19 in Qatar were young with few comorbidities. Case fatality rate was very low (only 7 deaths among 5,685 infected persons). Severe and critical illness were associated with presence of hypertension or diabetes.

## Introduction

A cluster of patients with pneumonia of unknown etiology linked to a seafood wholesale market was first reported from Wuhan, China in December 2019.^1-4^ A novel coronavirus, SARS-CoV-2 was isolated as the causative organism and the resultant disease was named COVID-19.^1,5^ Initially linked to the seafood market and presumed to be transmitted from animals to humans, the virus has since spread quickly across the globe through human-to-human transmission.^6-9^ As of May 3, 2020, more than 3.4 million cases and over 244,000 deaths have been reported globally. Published epidemiologic studies across a number of populations show significant differences in rates and severity of infection and in case fatality rates.^10^ At this stage of a novel virus pandemic, analyzing transmission patterns in populations with unique demographic characteristics can add to our understanding of the disease dynamics. While it is difficult to isolate the effects of public health measures, such as quarantine, lockdown, and physical distancing, it is nevertheless useful to track the course of the epidemic in relation to the timeline of their implementation. We describe the demographic characteristics, comorbidity profile, and disease severity of patients with confirmed COVID-19 infection in Qatar. We also report on significant public health measures implemented to slow the progression of the epidemic in Qatar.

Qatar is a part of the six-country Gulf Cooperation Council, which also includes Saudi Arabia, Kuwait, Oman, Bahrain and the United Arab Emirates. Qatar has a unique population demographic profile. Among the 2.8 million residents of Qatar, expatriate workforce constitutes about 88% of the population.^11^ Due to the nature of the expatriate workforce, the majority of the population in Qatar (∼75%) are male, and the population pyramid is heavily concentrated in the 20-50 year age groups, particularly among males.^11^ There is evidence that COVID-19 disproportionally affects males and outcomes are poorer in the older age group.^3,12,13^ Understanding the epidemiologic curve and risk factors for serious infection in Qatar will be important in understanding the epidemiology in countries with unique demographic characteristics.

## Methods

Even before the first case of COVID-19 was identified in the country, Qatar had instituted extensive plans to identify and manage persons with COVID-19 infection. The existing tracking, tracing and identification mechanism within the Ministry of Public Health, with proven effectiveness during the MERS-CoV outbreak, was expanded and put on alert.^14,15^ Testing for suspected cases started on February 5, 2020, and the first case was recorded on February 28, 2020. We used the tracking and reporting data from the Ministry of Public Health to determine the number of new cases diagnosed per day and their demographic characteristics between February 28, 2020 (date of identification of first case in Qatar) and April 18, 2020 (11:00 AM local time). All COVID-19 testing in Qatar was performed at the central laboratory of Hamad Medical Corporation, which is the public healthcare delivery arm for the State of Qatar and provides over 85% of the inpatient bed capacity in the State. Nasopharyngeal and throat swabs were collected from suspected cases with symptoms of influenza-like illness suggesting COVID- 19 and, if confirmed, from close contacts. Real time RT-PCR was used to detect SARS-CoV-2 infection. Nationality of each tested person was ascertained from the official State Identification Card, which is issued to each national and expatriate worker and their dependents residing in Qatar. Comorbidities were retrieved from the electronic medical records where they are coded using the International Classification of Diseases 10^th^ edition, Australian Modification. Severity of illness at the time of presentation was determined by expert coders using criteria suggested by the World Health Organization, including admission to an acute care or an intensive care bed, need for mechanical ventilation, oxygen saturation and supplemental oxygen requirement.^16^ **(Supplementary table 1)** Severity of illness was categorized into 1) asymptomatic or minimal symptoms, 2) mild symptoms or uncomplicated upper respiratory tract infection without clinical or radiographic evidence of pneumonia, 3) mild symptoms with clinical or radiographic evidence of pneumonia, 4) severely ill, and 5) critically ill.^16^

We created a timeline of newly diagnosed cases to study the progression of the epidemic in Qatar. Key governmental decisions taken by the Supreme Committee for Crisis Management and the Council of Ministers in response to the epidemic were marked on the timeline to show their temporal relation to the cases. Multivariable logistic regression was used to determine factors associated with severe and critical illness. Covariates of interest included in the model were age, gender, nationality and presence of comorbidities. Comorbidities with a total count of less than 15 were excluded because of the small numbers.

Changes in population movement was assessed using Google mobility reports, a publicly available tool that tracks movement of people who use mobile applications like Google Maps.^17^ They show changes in visits and length of stay at various locations compared to a baseline. Baseline was the median value, for the corresponding day of the week, during the 5-week period between January 3, 2020 and February 6, 2020.^17^

### Ethical Approval

The Institutional Review Board at Hamad Medical Corporation approved this study with an expedited status due to the emergency pandemic status of the COVID-19 outbreak.

### Patient and Public Involvement

This study was conducted in response to a national and global public health emergency. There was no patient or public involvement. However, key elements of the data are shared with the public on a daily basis.

## Results

Between February 5, 2020 and April 18, 2020 (11:00 AM local time), 60,645 persons were tested for SARS-CoV-2, of which 5,685 were confirmed positive. Before the first case was diagnosed, testing for SARS-CoV-2 focused on those with influenza-like illness and severe acute respiratory infection. The first cases were identified among quarantined travelers returning to Qatar on February 28, 2020, followed by the identification of a large cluster of over 300 infections on March 6, 2020 among expatriate craft and manual workers. Following the discovery of the first community cluster, testing was expanded to include contacts of new cases, persons in hotspots, that is areas where infections were diagnosed, individuals with suspected infection or suggestive symptoms, and travelers coming or returning to Doha. The rapid expansion of testing created a backlog, which was resolved through an investment in testing infrastructure that significantly increased the testing capacity to approximately 4,000 tests per day.

The number of new cases diagnosed by date of diagnosis is presented in **figure 1**. The epidemiologic curve showed two distinct patterns of infection transmission. A larger and sustained community transmission was observed among expatriate workers, predominantly among craft and manual workers, which subsequently reached other population segments. A second smaller cluster among Qatari nationals returning from overseas during the study period was not sustained over time. Subsequent smaller case clusters among Qatari nationals were traced to the local community. The number of tests performed per day and the number testing positive is provided in **supplementary figure 1**. The positivity rate, that is number of tests positive over total number of tests, is shown in **figure 2**. The positivity rate increased steadily with time, with somewhat of an accelerated rate after April 5, 2020.

**Figure 1.**
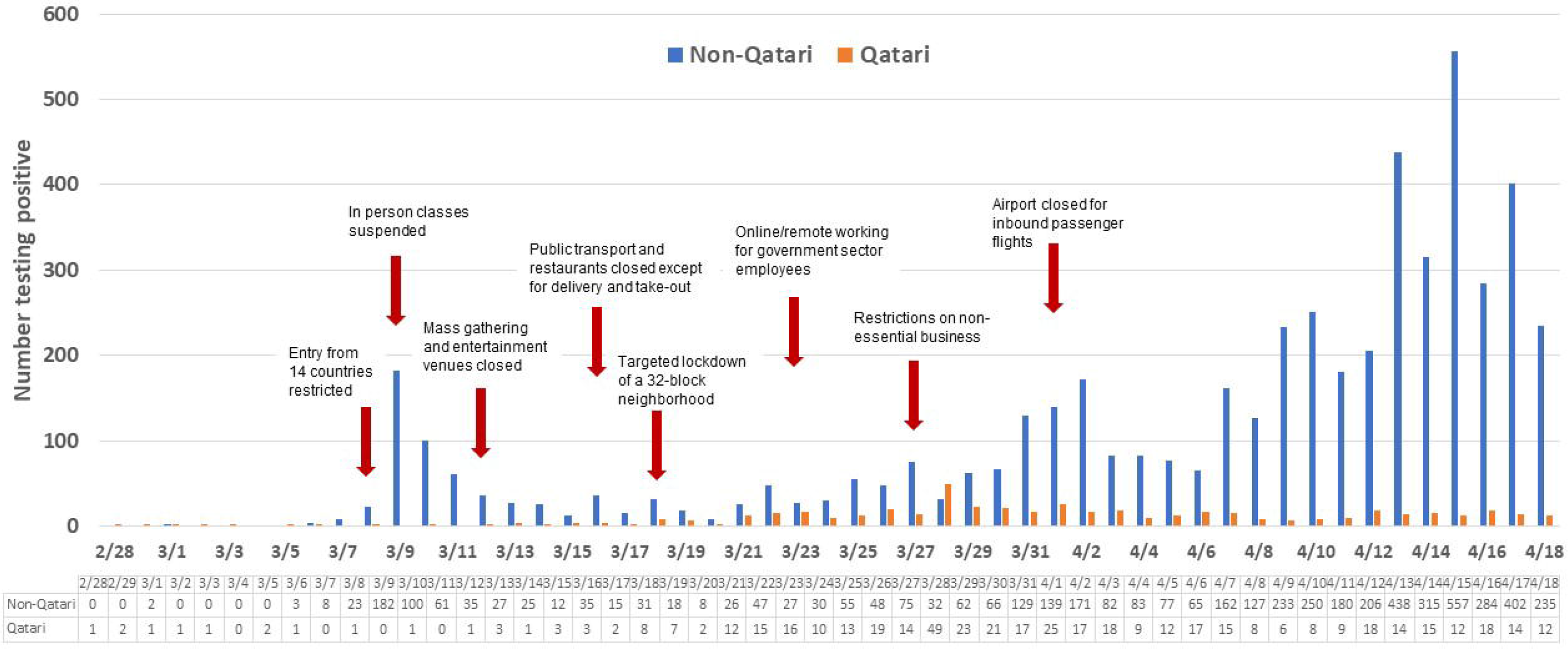
Epidemiologic curve of patients with COVID-19 in Qatar.

**Figure 2.**
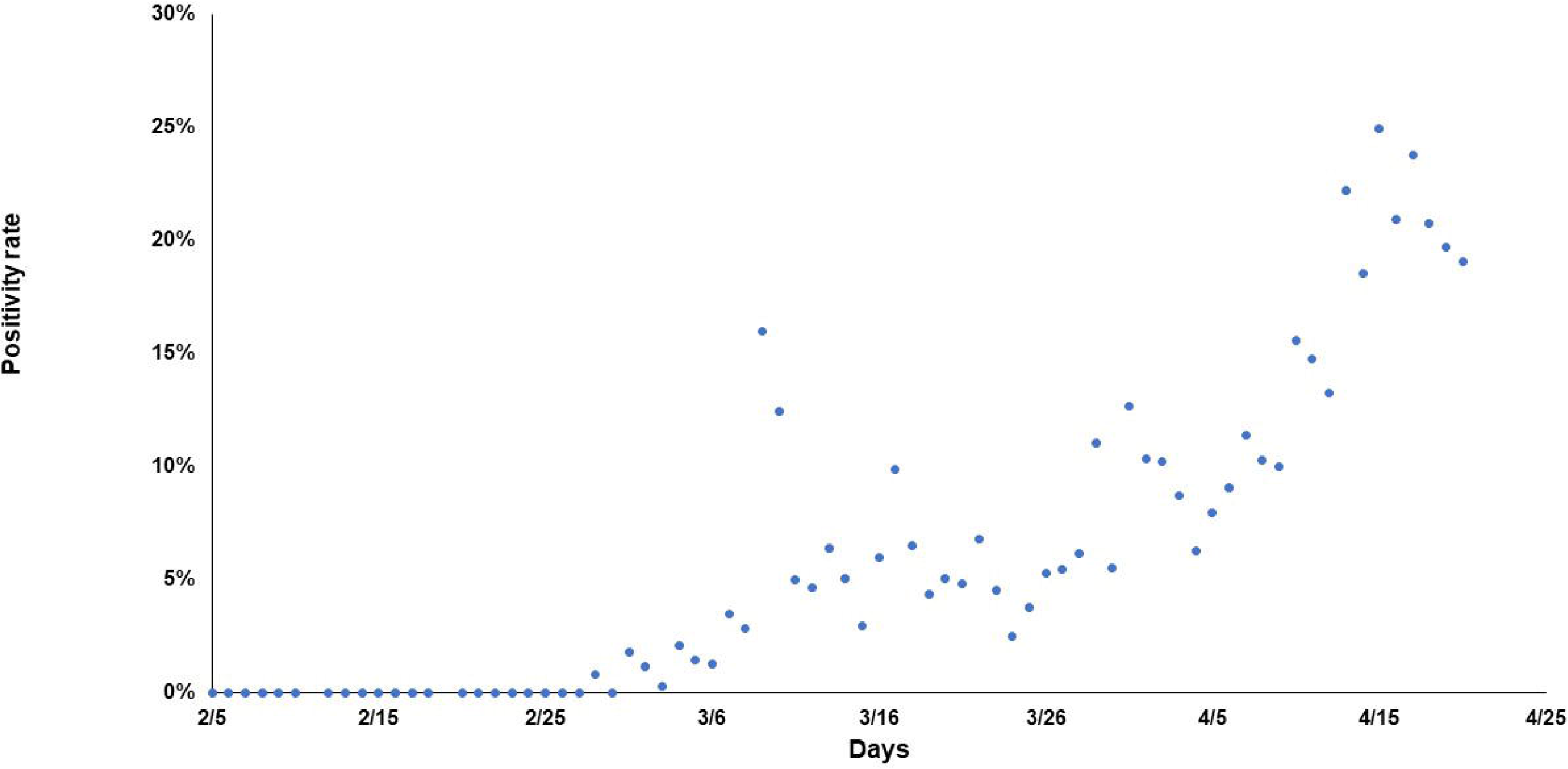
Positivity rate among those tested for SARS-CoV-2 active infection by swab day.

The mean age (SD) of the infected persons was 35.8 (12.0) years and 88.9% were male **(Table 1)**. Other baseline characteristics are also shown in **table 1**. The nationalities with highest frequency of infection were Indian (27.4%), Bangladeshi (18.9%), Nepalese (18.4%), Qatari (8.7%), and Pakistani (6.2%). The most common comorbidities were hypertension (6.9%), diabetes mellitus (6.0%), cardiovascular disease (4.4%) and chronic lung disease (3.0%). Comorbidity data were missing for 235 persons (4.1%). Among all infected persons, 4,753 (83.6%) had no known comorbidity and 697 (12.3%) had at least one comorbidity. An overwhelming majority of infected persons (90.9%) were either asymptomatic or had minimal symptoms, 0.8% had mild illness without evidence of pneumonia, 2.3% had mild illness with pneumonia, and 2% were severe or critically ill. Severity of illness data were missing for 223 (3.9%) persons. **(Table 1)**

**Table 1.**
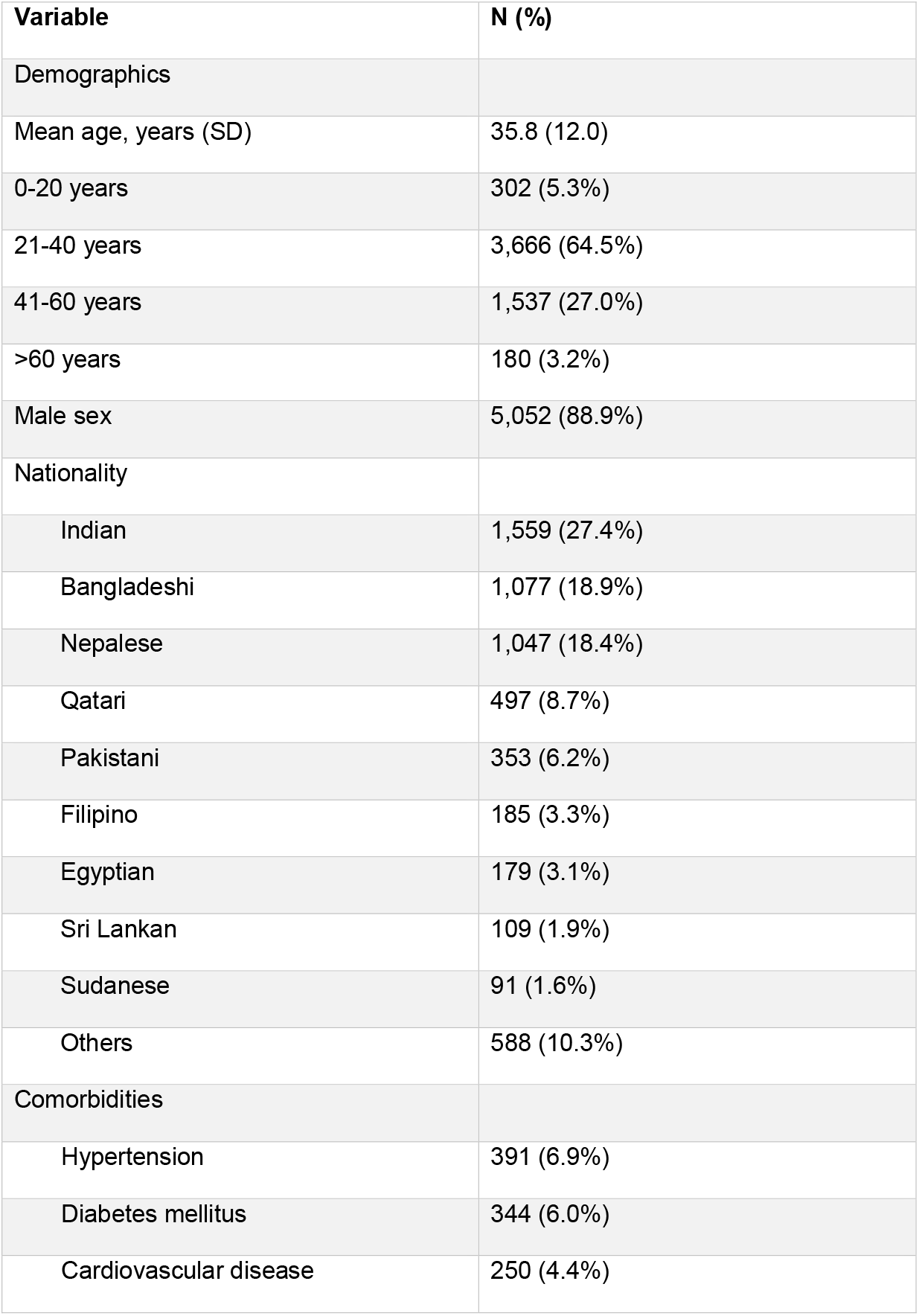
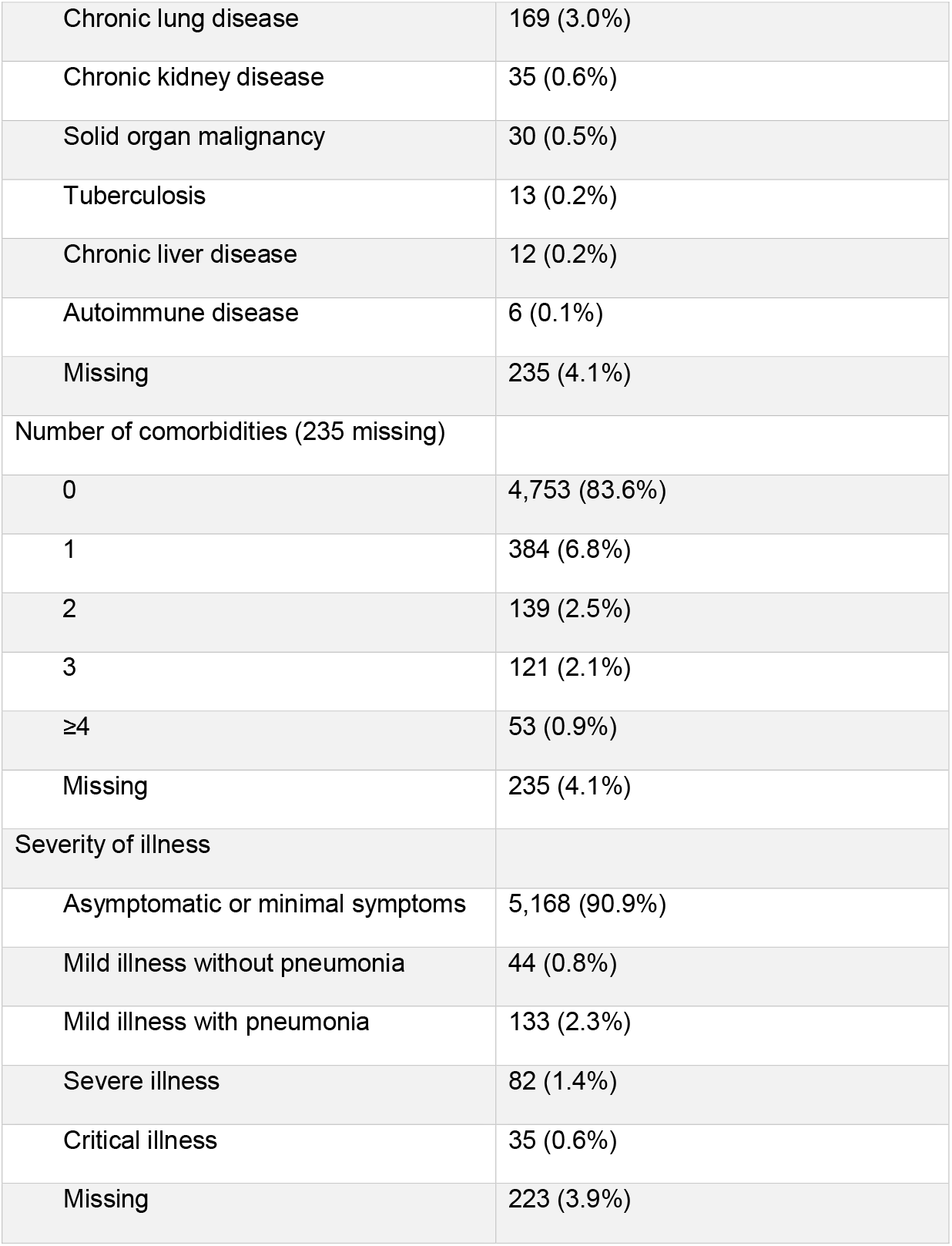
Characteristics of 5,685 patients with confirmed COVID-19 infection in Qatar between February 29 and April 18, 2020.

| Variable | N (%) |
| --- | --- |
| Demographics |  |
| Mean age, years (SD) | 35.8 (12.0) |
| 0-20 years | 302 (5.3%) |
| 21-40 years | 3,666 (64.5%) |
| 41-60 years | 1,537 (27.0%) |
| >60 years | 180 (3.2%) |
| Male sex | 5,052 (88.9%) |
| Nationality |  |
| Indian | 1,559 (27.4%) |
| Bangladeshi | 1,077 (18.9%) |
| Nepalese | 1,047 (18.4%) |
| Qatari | 497 (8.7%) |
| Pakistani | 353 (6.2%) |
| Filipino | 185 (3.3%) |
| Egyptian | 179 (3.1%) |
| Sri Lankan | 109 (1.9%) |
| Sudanese | 91 (1.6%) |
| Others | 588 (10.3%) |
| Comorbidities |  |
| Hypertension | 391 (6.9%) |
| Diabetes mellitus | 344 (6.0%) |
| Cardiovascular disease | 250 (4.4%) |
| Chronic lung disease | 169 (3.0%) |
| Chronic kidney disease | 35 (0.6%) |
| Solid organ malignancy | 30 (0.5%) |
| Tuberculosis | 13 (0.2%) |
| Chronic liver disease | 12 (0.2%) |
| Autoimmune disease | 6 (0.1%) |
| Missing | 235 (4.1%) |
| Number of comorbidities (235 missing) |  |
| 0 | 4,753 (83.6%) |
| 1 | 384 (6.8%) |
| 2 | 139 (2.5%) |
| 3 | 121 (2.1%) |
| ≥4 | 53 (0.9%) |
| Missing | 235 (4.1%) |
| Severity of illness |  |
| Asymptomatic or minimal symptoms | 5,168 (90.9%) |
| Mild illness without pneumonia | 44 (0.8%) |
| Mild illness with pneumonia | 133 (2.3%) |
| Severe illness | 82 (1.4%) |
| Critical illness | 35 (0.6%) |
| Missing | 223 (3.9%) |

A larger proportion of persons with mild illness with or without pneumonia and those with severe or critical illness had at least one comorbidity. **(Table 2)** Compared to those with asymptomatic or minimally symptomatic illness, prevalence of most comorbidities was 3-4 times higher among those with mild disease with or without pneumonia or severe/critical illness. Number of comorbidities by severity of illness is shown in **table 3**. Among persons with no comorbidity, 96.1% were asymptomatic or had minimal symptoms, 2.5% had mild illness with or without pneumonia, and 1.4% were severely or critical ill. Among persons with 2 or more comorbidities, 82.1% were asymptomatic or had minimal symptoms, 10.9% had mild illness with or without pneumonia, and 7.1% were severely or critical ill. **(Table 3)**

**Table 2.** Prevalence of comorbidities by severity of illness.

|  | **Asymptomatic or minimally symptomatic** | **Mild illness with or without pneumonia** | **Severe/critical illness** |
| --- | --- | --- | --- |
|  | **N (%)** | **N (%)** | **N (%)** |
| Hypertension | 317 (6.1%) | 41 (23.2%) | 31 (26.5%) |
| Diabetes mellitus | 281 (5.4%) | 33 (18.6%) | 30 (25.6%) |
| Cardiovascular disease | 199 (3.8%) | 31 (17.5%) | 19 (16.2%) |
| Chronic lung disease | 145 (2.8%) | 12 (6.8%) | 10 (8.6%) |
| Chronic kidney disease | 25 (0.5%) | 6 (3.4%) | 4 (3.4%) |
| Solid organ malignancy | 26 (0.5%) | 2 (1.1%) | 2 (1.7%) |
| Tuberculosis | 10 (0.2%) | 2 (1.1%) | 1 (0.8%) |
| Chronic liver disease | 10 (0.2%) | 2 (1.1%) | 0 (0%) |
| Autoimmune disease | 5 (0.1%) | 1 (0.6%) | 0 (0%) |

**Table 3.** Number of comorbidities by severity of illness.

| **Number of comorbidities** | **Asymptomatic or minimal**<br>**symptoms, N (%)** | **Mild illness with or without**<br>**pneumonia, N (%)** | **Severe/critical illness, N (%)** |
| --- | --- | --- | --- |
| No comorbidity | 4,582 (96.1%) | 120 (2.5%) | 67 (1.4%) |
| Only 1 comorbidity | 330 (86.6%) | 23 (6.0%) | 28 (7.3%) |
| 2 or more comorbidities | 256 (82.1%) | 34 (10.9%) | 22 (7.1%) |

Seven deaths were observed during the time interval studied, corresponding to a case-fatality rate of 1.2 per 1000 cases. All seven deaths were males aged 40-88 years. All except one (74 years old male) had comorbidities, including diabetes (5 subjects), cardiovascular disease (5 subjects) and hypertension (3 subjects). One patient, 58 years old had 5 comorbidities (diabetes, hypertension, cardiovascular disease, chronic kidney and liver disease). In a multivariable logistic regression model, presence of hypertension (OR 3.49; 95% CI 1.83,6.68) or diabetes (OR 3.17; 95% CI 1.76,5.71) were associated with a higher risk of severe or critical disease. **(Table 4)** Cardiovascular disease, chronic lung disease, chronic kidney disease and solid organ malignancy were not associated with a higher risk. We repeated the logistic regression analysis after excluding those with missing data and the results were nearly identical. **(Supplementary table 2)** We also repeated the analysis using number of comorbidities as covariates. Presence of any single comorbidity (OR 5.43, 95% CI 3.41,8.63) or any 3 or more comorbidities (OR 6.16, 95% CI 3.35,11.32) were associated with a higher risk of severe or critical illness. **(Supplementary table 3)**

**Table 4.**
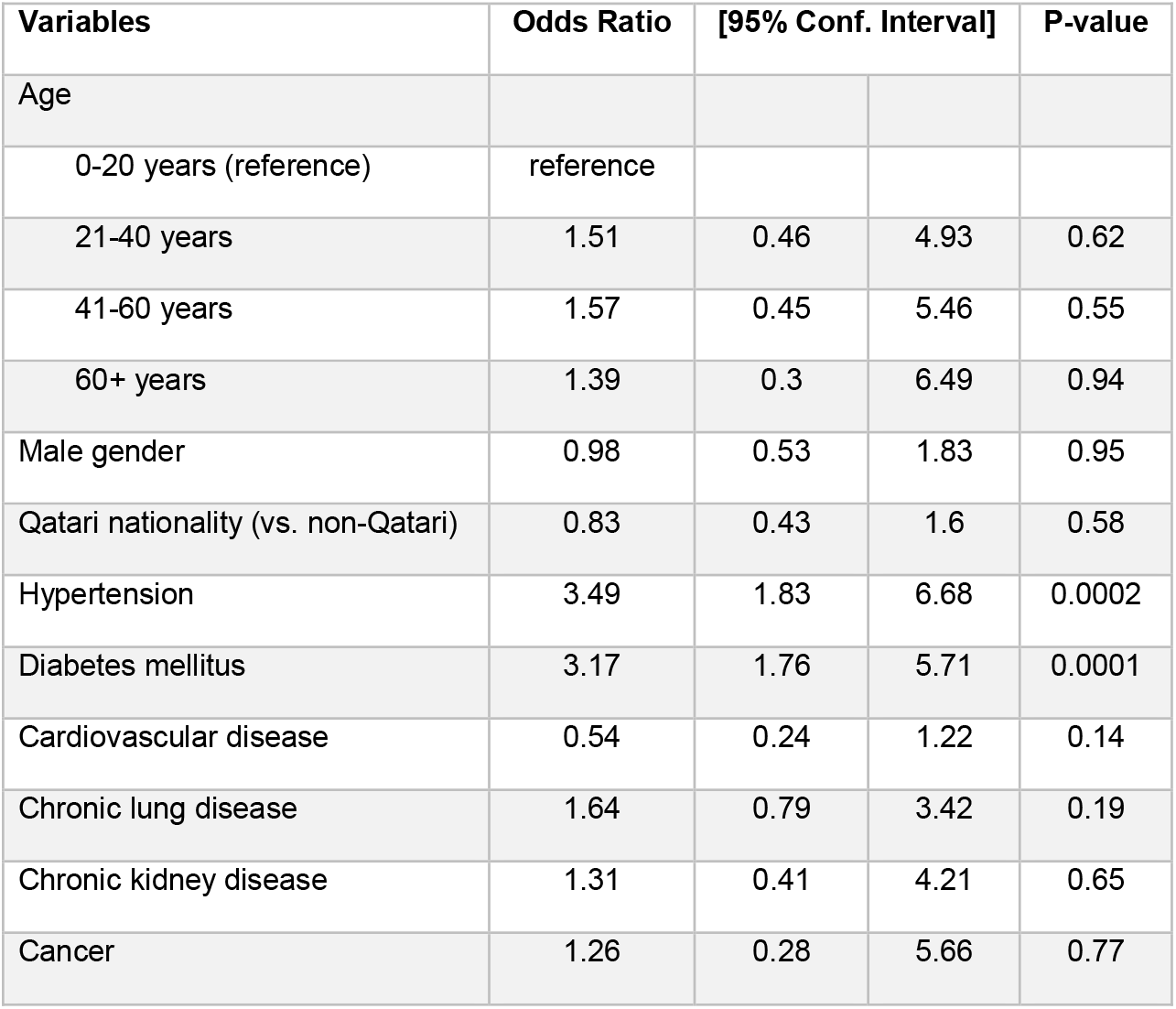
Factors associated with severe or critical illness (multivariable logistic regression model).

| Variables | Odds Ratio | [95% Conf. Interval] |  | P-value |
| --- | --- | --- | --- | --- |
| Age |  |  |  |  |
| 0-20 years (reference) | reference |  |  |  |
| 21-40 years | 1.51 | 0.46 | 4.93 | 0.62 |
| 41-60 years | 1.57 | 0.45 | 5.46 | 0.55 |
| 60+ years | 1.39 | 0.3 | 6.49 | 0.94 |
| Male gender | 0.98 | 0.53 | 1.83 | 0.95 |
| Qatari nationality (vs. non-Qatari) | 0.83 | 0.43 | 1.6 | 0.58 |
| Hypertension | 3.49 | 1.83 | 6.68 | 0.0002 |
| Diabetes mellitus | 3.17 | 1.76 | 5.71 | 0.0001 |
| Cardiovascular disease | 0.54 | 0.24 | 1.22 | 0.14 |
| Chronic lung disease | 1.64 | 0.79 | 3.42 | 0.19 |
| Chronic kidney disease | 1.31 | 0.41 | 4.21 | 0.65 |
| Cancer | 1.26 | 0.28 | 5.66 | 0.77 |

Google mobility reports data demonstrated a significant decrease in number of people visiting retail and recreation outlets, grocery and pharmacy stores, parks, transit stations and workplaces over time. **(Table 5)** A concurrent increase in people staying in residential areas was observed over this timeframe. A snapshot on April 17 shows a 69% reduction in visits to retail and recreation areas, a 44% reduction in visits to grocery and pharmacy stores, a 64% decrease in visits to parks, a 68% decrease in visits to transit stations and a 38% decrease in visits to workplaces. A 21% increase in people at residential areas was observed on this date.

**Table 5.**
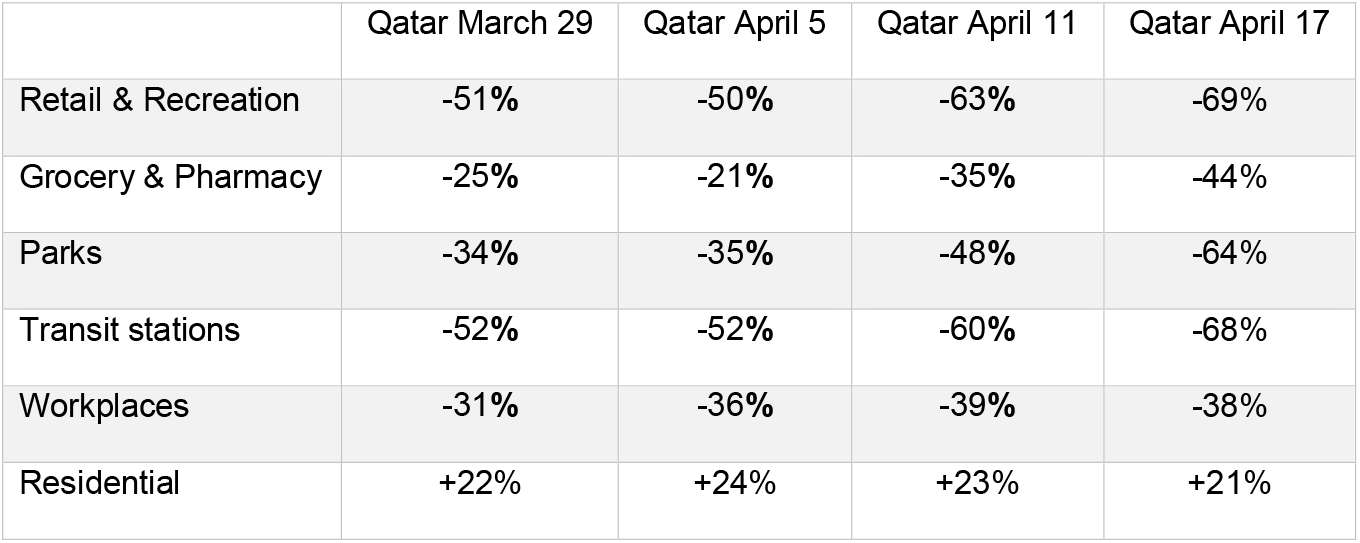
Relative change in people visiting areas of common interest in the State of Qatar. (Comparison period January 3, 2020 – February 6, 2020)

## Discussion

We provide a characterization of the SARS-CoV-2 outbreak in Qatar, which offers new insights into the behavior of the pandemic in a unique demographic setting.

The first case in Qatar was identified on February 28, 2020, among returning travelers, which is nearly 9 weeks after the first cluster was reported from Wuhan, China.^18^ During this time, infection had spread to multiple countries over four continents. This was also a critical time during which the State of Qatar formulated a national plan to respond to the anticipated cases. Testing for SARS-CoV-2 started in Qatar on February 5, 2020 and the first major cluster of cases was identified on March 8, 2020 where over 300 cases were linked to 4 expatriate workers through aggressive contact tracing. Such aggressive identification and contact tracing and testing were probably the reasons for a small number of daily new cases till March 31, 2020. At that time, a large number of returning travelers and nationals were identified to have infection. The number of daily diagnosed infections accelerated in April, in part due to a large increase in number of daily tests, but also reflecting expansion of the epidemic in the wider population. This trend occurred predominantly in expatriate workers often living in more crowded areas and accommodations with frequent social mixing despite a national campaign to discourage peoples’ movement except in urgent situations. The epidemic, however, eventually reached a larger population.

An overwhelming majority (>90%) of confirmed COVID-19 cases were asymptomatic or with minimal symptoms not requiring urgent medical care or hospitalization. This is likely due to the younger age of the population (mean age 35.8 years) and overall absence of any comorbidities in the vast majority of the infected persons. This reinforces our current understanding of the disease being mild or asymptomatic in a majority of the persons, particularly among the younger and healthier persons, as well as the strong role of age in the epidemiology of this infection. Similar to other studies, presence of comorbidities was associated with severe or critical disease.^1^ We found a very low mortality among confirmed COVID-19 patients in Qatar, which may at least partly be attributable both to the timely and effective response of the health system and the demographic characteristics of the infected persons. It is conceivable that right censoring with the time delay between onset of disease to death may play a part, though similarly very low mortality after the study period ended does not support this. The role that factors such as free access to high quality medical care for everybody in Qatar, availability of a high number of critical care beds, or differences in viral subtypes, played, needs further study.

In response to the spread of COVID-19, the country took a series of public health measures, including limiting incoming passenger flights into Doha through Hamad International Airport and providing free state quarantine facilities for returning travelers. A host of other measures were implemented gradually that promoted physical distancing including, closing retail stores in malls and shopping centers, closing entertainment and dining facilities, postponing or canceling large sports events and conferences, suspending classes in schools and universities, and mandating working from home for 80% of workers in the public and private sectors. The healthcare system was also reorganized to prioritize COVID-19 response over routine services. All positive cases including those without symptoms were admitted to isolation facilities by the public healthcare system. These public health measures were heavily promoted and widely communicated through social and traditional media outlets to all segments of society. Mobility data shows a significant reduction in visits to common retail, recreation, transit and workplace areas, which may have contributed to a reduction in spread of infection. There was a more pronounced reduction in mobility in the weeks following April 5th, which coincides with the significant rise in the reported number of positive cases.

Strengths of our study include unified contact tracing and testing, with all testing done at a single lab. All tests performed in the State of Qatar were included, providing a robust national estimate of the number of infected persons among those tested. There are limitations to our study. Comorbidities were retrieved from the electronic medical records using ICD-10 AM codes. Exact geographic location and contact tracing data were not included in the current report. Our study end date was April 18, 2020, and all persons with confirmed infection till that date were included. However, it is possible that some persons may have progressed to more severe disease after this date.

In conclusion, we describe the evolution of COVID-19 epidemic in the State of Qatar. The epidemic predominantly affected males and younger population and was associated with no or minimal symptoms in a vast majority of the infected persons. Public health measures were instituted early and may have led to the slower growth compared with other countries which delayed such measures.

## Data Availability

None

## Competing interests

None of the authors have any financial conflict of interest related to this article.

H.E. Dr. Hanan. M. Al Kuwari is the Minister of Public Health for the State of Qatar.

## Ethical Approval

This study was approved by the Institutional Review Board at Hamad Medical Corporation. (MRC-05-011)

## Patient Consent

A waiver of informed consent was granted.

## Data Sharing Statement

No additional data are available

## Patient of Public Involvement

There was not patient or public involvement in the design, conduct or reporting of this study.

## Transparency declaration

The lead author (the manuscript’s guarantor) affirms that the manuscript is an honest, accurate, and transparent account of the study being reported; that no important aspects of the study have been omitted; and that any discrepancies from the study as planned (and, if relevant, registered) have been explained.

## Dissemination declaration

Dissemination to study participants or patient organizations is not possible/applicable.

